# Secondary infections modify the overall course of hospitalized COVID-19 patients: A retrospective study from a network of hospitals across North India

**DOI:** 10.1101/2021.09.27.21264070

**Authors:** Sandeep Budhiraja, Bansidhar Tarai, Dinesh Jain, Mona Aggarwal, Abhaya Indrayan, Poonam Das, RS Mishra, Supriya Bali, Monica Mahajan, Jay Kirtani, Rommel Tickoo, Pankaj Soni, Vivek Nangia, Ajay Lall, Nevin Kishore, Ashish Jain, Omender Singh, Namrita Singh, Ashok Kumar, Prashant Saxena, Arun Dewan, Ritesh Aggarwal, Mukesh Mehra, Meenakshi Jain, Vimal Nakra, B D Sharma, Praveen Kumar Pandey, YP Singh, Vijay Arora, Suchitra Jain, Ranjana Chhabra, Preeti Tuli, Vandana Boobna, Alok Joshi, Manoj Aggarwal, Rajiv Gupta, Pankaj Aneja, Sanjay Dhall, Vineet Arora, Inder Mohan Chugh, Sandeep Garg, Vikas Mittal, Ajay Gupta, Bikram Jyoti, Puneet Sharma, Pooja Bhasin, Shakti Jain, RK Singhal, Atul Bhasin, Anil Vardani, Vivek Pal, Deepak Gargi Pande, Tribhuvan Gulati, Sandeep Nayar, Sunny Kalra, Manish Garg, Rajesh Pande, Pradyut Bag, Arpit Gupta, Jitin Sharma, Anil Handoo, Purabi Burman, Ajay Kumar Gupta, Pankaj Nand Choudhary, Ashish Gupta, Puneet Gupta, Sharad Joshi, Nitesh Tayal, Manish Gupta, Anita Khanna, Sachin Kishore, Shailesh Sahay, Rajiv Dang, Neelima Mishra, Sunil Sekhri, Rajneesh Chandra Srivastava, Mitali Bharat Agrawal, Mohit Mathur, Akash Banwari, Sumit Khetarpal, Sachin Pandove, Deepak Bhasin, Harpal Singh, Devender Midha, Anjali Bhutani, Manpreet Kaur, Amarjit Singh, Shalini Sharma, Komal Singla, Pooja Gupta, Vinay Sagar, Ambrish Dixit, Rashmi Bajpai, Vaibhav Chachra, Puneet Tyagi, Sanjay Saxena, Bhupesh Uniyal, Shantanu Belwal, Imliwati Aier, Mini Singhal, Ankit Khaduri

## Abstract

**Introduction:** SARS-CoV-2 infection increases the risk of secondary bacterial and fungal infections and contributes to adverse outcomes. The present study was undertaken to get better insights into the extent of secondary bacterial and fungal infections in Indian hospitalized patients and to assess how these alter the course of COVID-19 so that the control measures can be suggested.

**Methods:** This is a retrospective, multicentre study where data of all RT-PCR positive COVID-19 patients was accessed from Electronic Health Records (EHR) of a network of 10 hospitals across 5 North Indian states, admitted during the period from March 2020 to July 2021.The data included demographic profile of patients, clinical characteristics, laboratory parameters, treatment modalities, and outcome in those with secondary infections (SIs) and those without SIs. Spectrum of SIS was also studied in detail.

**Results:** Of 19852 RT-PCR positive SARS-CO2 patients admitted during the study period, 1940 (9.8%) patients developed SIs. Patients with SIs were 8 years older on average (median age 62.6 years versus 54.3 years; P<0.001) than those without SIs. The risk of SIs was significantly (p < 0.001) associated with age, severity of disease at admission, diabetes, ICU admission, and ventilator use.

The most common site of infection was urinary tract infection (UTI) (41.7%), followed by blood stream infection (BSI) (30.8%), sputum/BAL/ET fluid (24.8%), and the least was pus/wound discharge (2.6%). As many as 13.4% had infections with more than organism and 34.1% patients had positive cultures from more than one site. Gram negative bacilli (GNB) were the commonest organisms (63.2%), followed by Gram positive cocci (GPC) (19.6%) and fungus (17.3%). Most of the patients with SIs were on multiple antimicrobials – the most commonly used were the BL-BLI for GNBs (76.9%) followed by carbapenems (57.7%), cephalosporins (53.9%) and antibiotics carbapenem resistant entreobacteriace (47.1%). The usage of emperical antibiotics for GPCs was in 58.9% and of antifungals in 56.9% of cases, and substantially more than the results obtained by culture.

The average stay in hospital for patients with SIs was twice than those without SIs (median 13 days versus 7 days). The overall mortality in the group with SIs (40.3%) was more than 8 times of that in those without SIs (4.6%). Only 1.2% of SI patients with mild COVID-19 at presentation died, while 17.5% of those with moderate disease and 58.5% of those with severe COVID-19 died (P< 0.001). The mortality was highest in those with BSI (49.8%), closely followed by those with HAP (47.9%), and then UTI and SSTI (29.4% each). The mortality rate where only one microorganism was identified was 37.8% and rose to 56.3% in those with more than one microorganism. The mortality in cases with only one site of infection was 28.8%, which steeply rose to 62.5% in cases with multiple sites of infection. The mortality in diabetic patients with SIs was 45.2% while in non-diabetics it was 34.3% (p < 0.001).

**Conclusions:** Secondary bacterial and fungal infections can complicate the course of almost 10% of COVID-19 hospitalised patients. These patients tend to not only have a much longer stay in hospital, but also a higher requirement for oxygen and ICU care. The mortality in this group rises steeply by as much as 8 times. The group most vulnerable to this complication are those with more severe COVID-19 illness, elderly, and diabetic patients. Varying results in different studies suggest that a region or country specific guideline be developed for appropriate use of antibiotics and antifungals to prevent their overuse in such cases. Judicious empiric use of combination antimicrobials in this set of vulnerable COVID-19 patients can save lives.

## Introduction

Viral infections, particularly SARS-Co-V-2, may predispose to concomitant and subsequent bacterial infections [1,2]. Various explanations given for this phenomenon include direct damage to the respiratory epithelium caused by the virus, their effects on innate and adaptive immunity, and SARS-CoV-2 associated perturbation of gut homeostasis [1,2].

Secondary bacterial infections have been noted to be a significant contributor to increased morbidity and mortality in earlier influenza pandemics and during seasonal influenza, and also in other respiratory diseases [3,4]. Shafran et al. [5] found that COVID-19 patients had a higher rate of secondary bacterial infections compared to influenza patients (12.6% versus 8.7%, p=0.006). Other studies suggested that superinfections, especially in the later stage of illness, were encountered in 8% of patients with COVID-19, usually those that were more severely ill and those who died [5,6,7,8].

Because of concerns of increased mortality in patients with bacterial superinfections during influenza pandemics, several guidelines advocate the use of empirical antibiotics for patients with severe COVID-19 [9,10]. This, however, has a potential of antibiotic overuse and increasing antimicrobial resistance [11]. As the prevalence of secondary bacterial and fungal infections in COVID-19 patients in India is not clearly known, a better understanding would be crucial for treating COVID-19 and to help ensure responsible use of antimicrobials to minimize negative consequences of overuse.

The present study was undertaken to get better insights into the extent of secondary bacterial and fungal infections in Indian patients of COVID-19 admitted to hospitals and to assess how these alter the course of the disease. Evaluating the treatment strategies used in this group of patients may help design appropriate guidelines for empirical use of antimicrobials in COVID-19 Indian patients.

## Methods

This is a retrospective, multicentre study where data of all RT-PCR positive COVID-19 patients was accessed from Electronic Health Records (EHR) of a network of 10 hospitals across 5 North Indian states, admitted during the period from March 2020 to July 2021. They were divided in to mild, moderate and severe categories as per the government of India criteria [12]. The data included demographic profile of patients, presence of diabetes, various investigations like CRP, D-Dimer, IL-6, ferritin, CPK, LDH, Trop-I and lymphocyte counts, wherever available, the HRCT chest severity score (CTSS), various treatment modalities like use of steroids, Remdesivir and convalescent plasma, average length of stay and in-hospital mortality.

Detailed data were available for secondary infections (SIs). The microbiological data in the form of culture results from blood, urine, pus/wound discharge, sputum, BAL fluid culture, and ET secretion cultures, was analysed and patients were categorized into four types of Sis, namely, blood stream infection (BSI), urinary tract infection (UTI), skin and soft tissue infection (SSTI) and hospital acquired pneumonia (HAP). The patients with SIs were compared with those without SIs for these parameters. Use of antibiotics, antifungals, and antivirals in those who developed secondary infections (SIs) was studied. Substantial number of patients was on multiple antimicrobials, and many had multiple sites of infections. For statistical analysis, these were included under one predominant or primary site of infection if the same organism was isolated from different sites. Those who had more than two sites involved with same micro-organism were categorised to have clinically primary site of infection, as clinically evident or supported by radiological evidence.

### Statistical Analysis

The data have been presented as counts and percentages for qualitative characteristics such as sex, place of admission, and use of oxygen, and as mean and SD for quantitative characteristics such as age. Length of stay in the hospital and laboratory parameters have been summarised in terms of median and inter-quartile rang (IQR) because of their highly skewed distribution. Statistical significance of the difference between cases with SI and without SI was assessed by chi-square test or Student t-test. Fisher exact test was used for comparing small (<5) frequencies. For highly skewed distributions, such as of inflammatory markers, Wilcoxon-Mann-Whitney test was used. A p-value less than 0.05 was considered statistically significant although, in this case, the number of cases is so large for some categories that p-values have to be cautiously interpreted. SPSS 21 was used for calculations.

### Ethics Committee Approval and Consent

The study was approved by the Institutional Ethics Committee, Max Super Speciality Hospital (A unit of Devki Devi Foundation), Address: Service Floor, Office of Ethics Committee, East Block, next to Conference Room, Max Super Speciality Hospital, Saket (A unit of Devki Devi Foundation), 2, Press Enclave Road, Saket, New Delhi – 110017 vide ref. no. BHR/RS/MSSH/DDF/SKT-2/IEC/IM/21-24 dated 7th September 2021. The IEC provided no objection and approved the publication of this manuscript.

All the admitted patients gave a prior consent for their anonymised data to be used for research purposes.

## Results

### Demographics

A total of 19852 RT-PCR positive SARS-COV2 patients were admitted during the period of the study in the network of our hospitals. Their records were retrieved from the electronic health record system. Of these, a total of 1940 (9.8%) patients developed secondary infections. No significant (p = 0.100) gender difference was observed but the patients with SIs were on average 8 years older than those without secondary infection (median age 62.6 years versus 54.3 years; p < 0.001) (Table 1). As the age increased, the incidence of SIs steeply increased from 4.0% in <45 years to 18.4% in ≥75 years. More than one-fifth (22.6%) of ICU patients were affected against only 3.0% ward patients (p < 0.001). Those on oxygen were significantly (p < 0.0010) more affected (12.1%) than those not on oxygen (4.9%) and the incidence increased with the increasing need of oxygen from O2 to non-invasive ventilation (NIV) to mechanical ventilation (MV). Such details for those on convalescent plasma therapy (CPT), steroids, and remdesivir are also given in Table 1. The median length of stay was almost double (13 days) in the SI group compared to the group without SIs (7 days, p <0.001) (Table 1).

**Table-1:** Comparison of characteristics of cases with and without infection.

| Parameters |  | Total cases |  | Cases with SIs |  | Cases without SIs |  | P-value* |
| --- | --- | --- | --- | --- | --- | --- | --- | --- |
|  |  | Number | Percent of total | Number | Percent infected | Number | Percent not infected |  |
| Total |  | 19852 | 100.0 | 1940 | 9.8 | 17912 | 90.2 | X |
| Sex | Male | 13175 | 66.4 | 1255 | 9.5 | 11920 | 90.5 | 0.100 |
|  | Female | 6677 | 33.6 | 685 | 10.3 | 5992 | 89.7 |  |
| Age (Years) | <45 | 5547 | 27.9 | 222 | 4.0 | 5325 | 96.0 | <0.001 |
|  | 45 – 59 | 6369 | 32.1 | 523 | 8.2 | 5846 | 91.8 |  |
|  | 60 – 74 | 6112 | 30.8 | 860 | 14.1 | 5252 | 85.9 |  |
| | $\geq 75$ | 1824 | 9.2 | 335 | 18.4 | 1489 | 81.6 | |
| Mean age (years) (SD) |  | 55.0 (15.9) |  | 62.6 (13.9) |  | 54.4 (15.9) |  | <0.001 |
| Admission | ICU | 6845 | 34.5 | 1548 | 22.6 | 5297 | 77.4 | <0.001 |
|  | Ward | 13007 | 65.5 | 392 | 3.0 | 12615 | 97.0 |  |
| Oxygen | No | 6402 | 32.2 | 312 | 4.9 | 6090 | 95.1 | <0.001 |
|  | Yes | 13450 | 67.8 | 1628 | 12.1 | 11822 | 87.9 |  |
| Type of oxygen (percentage out of those who received oxygen) | O2 (NP/FM/NRBM) | 9842 | 49.6 | 822 | 8.4 | 9020 | 91.6 | <0.001 |
|  | NIV | 1800 | 9.1 | 390 | 21.7 | 1410 | 78.3 |  |
|  | MV | 1808 | 9.1 | 416 | 23.0 | 1392 | 77.0 |  |
| CPT | Given | 2542 | 12.8 | 685 | 26.9 | 1857 | 73.1 | <0.001 |
|  | Not Given | 17310 | 87.2 | 1255 | 7.3 | 16055 | 92.7 |  |
| Steroids (Drugs) | Given | 18344 | 92.4 | 1893 | 10.3 | 16451 | 89.7 | <0.001 |
|  | Not Given | 1508 | 7.6 | 47 | 3.1 | 1461 | 96.9 |  |
| Remdesivir | Given | 12017 | 60.5 | 1273 | 10.6 | 10744 | 89.4 | <0.001 |
|  | Not Given | 7835 | 39.5 | 667 | 8.5 | 7168 | 91.5 |  |
| Length of stay (days) | Median (IQR) | 7 (5.12-10.14) |  | 13 (7.87-20.05) |  | 7 (5.05-9.86) |  | <0.001 |

### Characteristics of SIs

Of the 1940 patients with SIs, 598 (30.8%) had positive blood culture, 809 (41.7%) had positive urine culture, 51 (2.6%) had positive cultures from pus / wound discharge, and 482 (24.8%) had positive cultures from sputum / BAL fluid or ET secretions (Table2). Thus, the most common site was urine, followed by blood. Mean age of patients with infections at different sites was not significantly different (p = 0.315). Of all infections, HAP was significantly less in females (18.5%) compared to males (28.3%) whereas UTI was significantly more (48.9% vs. 37.8%, p < 0.0010). Those with diabetes had relatively more of BSI and UTI and less of SSTI and HAP.

**Table 2:** Comparison of characteristics of cases with SIs at different sites (n = 1940)

| Parameters |  | BSI |  | UTI |  | SSTI |  | HAP |  | P-value |
| --- | --- | --- | --- | --- | --- | --- | --- | --- | --- | --- |
|  |  | Number | Percent | Number | Percent | Number | Percent | Number | Percent |  |
| Total – Percent out of total COVID cases |  | 598 | 3.0 | 809 | 4.1 | 51 | 0.3 | 482 | 2.4 |  |
| Total – Percent out of the cases with SIs |  | 598 | 30.8 | 809 | 41.7 | 51 | 2.6 | 482 | 24.8 |  |
| Sex | Male | 395 | 31.5 | 474 | 37.8 | 31 | 2.5 | 355 | 28.3 | <0.001 |
|  | Female | 203 | 29.6 | 335 | 48.9 | 20 | 2.9 | 127 | 18.5 |  |
| Age (Years) | Mean (SD) | 62.6 (13.8) |  | 62.9 (14.1) |  | 61.3 (15.0) |  | 62.4 (13.5) |  | 0.315 |
| Diabetes | Yes | 350 | 32.8 | 463 | 43.4 | 14 | 1.3 | 239 | 22.4 | <0.001 |
|  | No | 248 | 28.4 | 346 | 39.6 | 37 | 4.2 | 243 | 27.8 |  |

A significant number of patients had infection with more than one organism (261 patients out of 1940; 13.4%) and 662 patients (34.1%) had positive cultures from more than one site. Overall, there were 685 positive cultures in blood (80.4% bacterial and 19.6% fungal), 893 positive cultures in urine (72.4% bacterial and 27.6% fungal), 607 positive cultures from any of the respiratory secretions (99.7% bacterial and only 2 samples positive for Candida auris), and 63 positive cultures from pus / wound discharge (93.7% bacterial and 6.3% fungal). Species wise details are in Figure 1. Overall, there were 2248 positive isolates (bacterial and fungal) from samples of 1940 patients who had SIs. Of these, 1420 (63.2%) were Gram negative bacilli (GNB), 440 (19.6%) Gram positive cocci (GPC), and 388 (17.3%) fungal (Candida sp.). The breakup of Candida species in blood and urine is shown in Table 3. Candida albicans and Candida tropicalis were more common in urine and Candida auris in blood.

**Table3:** Breakup of Candida species in BSI and UTI.

| Candida species | BSI |  | UTI |  | P-value* |
| --- | --- | --- | --- | --- | --- |
|  | Count | Percent | Count | Percent |  |
| Candida albicans | 23 | 16.9 | 71 | 28.9 | 0.009 |
| Candida auris | 35 | 25.7 | 33 | 13.4 | 0.003 |
| Candida catenulate | 1 | 0.7 | 0 | 0 | 0.356 |
| Candida ciferrii | 1 | 0.7 | 0 | 0 | 0.356 |
| Candida famata | 3 | 2.2 | 0 | 0 | 0.044 |
| Candida duobushaemulonii | 0 | 0 | 5 | 2.0 | 0.165 |
| Candida glabrata | 15 | 11.0 | 17 | 6.9 | 0.164 |
| Candida guilliermondii | 1 | 0.7 | 1 | 0.4 | >0.999 |
| Candida kefyr | 1 | 0.7 | 0 | 0 | 0.356 |
| Candida krusei | 2 | 1.5 | 1 | 0.4 | 0.289 |
| Candida parapsilosis | 13 | 9.6 | 7 | 2.8 | 0.005 |
| Candida tropicalis | 41 | 30.1 | 111 | 45.1 | 0.004 |
| **Total** | **136** | **100.0** | **246** | **100.0** |  |

\*For comparison of each vs. the rest

**Figure 1:**
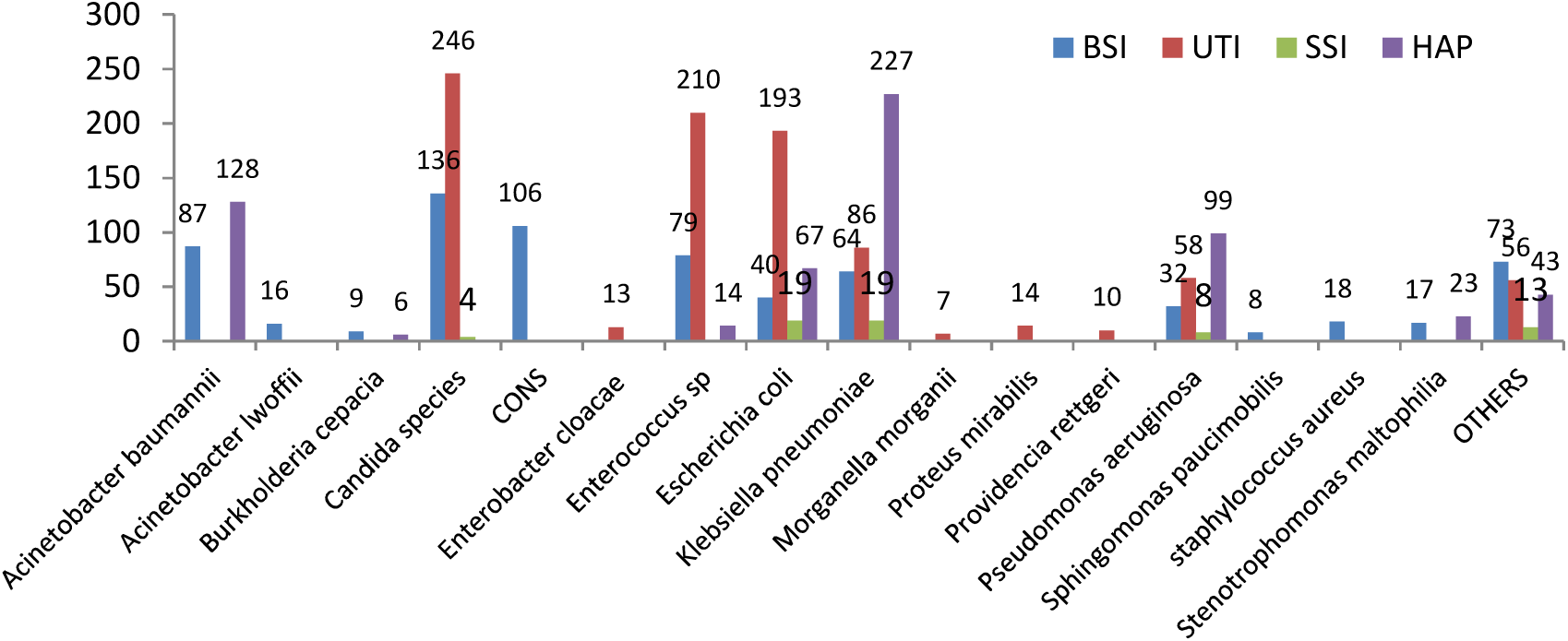
Breakup of microbiological flora causing SIs.

### Antimicrobial Resistance Pattern

Antibiotic sensitivity test was conducted among the samples tested for secondary infection. Carbapenem resistance was observed in 68% of *Acinetobacter baumanii*, 48% of *Klebseilla pneumonia*, 39% of *E. coli*, and 43% cases of *Pseudomonas aeruginosa*. Fluconazole resistance was studied in cultures positive for fungal infection and found fluconazole resistance in 54% of *Candida auris*, 10% of *Candida albicans*, and 19% of non-albicans Candida.

### Antimicrobial Treatment

The usage of various antimicrobial agents (antibiotics, antifungals, and antivirals) as initial empirical therapy in those patients who developed SIs was also studied. Almost all these patients were on multiple antibiotics and/or antifungals. In terms of usage of antimicrobials for COVID-19 infection, the commonly used medications were Remdesivir (74.8%), Favipiravir (21.2%), Doxycycline (50.2%), Ivermectin (43.5%) and Azithromycin (29.3%). For empirical treatment of SIs, the most used antibiotics were those directed against GNBs. The most used antibiotics against GNBs were BL-BLIs (76.9%), carbapenems (57.7%), cephalosporins (53.9%), and antibiotics against CREs (47.1%). Empirical usage of antibiotics against GPCs was seen in 58.9% of the patients with SIs. Interestingly, we observed empirical usage of antifungals in 56.9% of the patients with SIs. See Table 4 for details.

**Table 4:** Usage of various antimicrobial agents in SI group.

| Antimicrobial Agent | Sub Agent | Number of cases | Percent of total SI cases |
| --- | --- | --- | --- |
| Antibiotics for GNB CRE | Polymyxin B (589, 64.4%), Colistin (166, 18.2%), Fosfomycin (83, 9.1%), Minocycline (43, 4.7%), Tigicycline (33, 3.6%) | 914 | 47.1 |
| Carbapenems for GNB | Meropenem (949, 84.7%), Doripenem (42, 3.8%), Imipenem (20, 1.8%), Ertapenem (109, 9.7%) | 1120 | 57.7 |
| BL-BLI for GNB | Piperacillin-Tazobactam (846, 56.7%), Ceftriaxone-Sulbactam (373, 25%), Ticarcillin-Clavulanate (139, 9.3%), Others (133, 8.9%) | 1491 | 76.9 |
| Cephalosporins for GNB | Ceftriaxone (468, 44.8%), Cefepime (326, 31.2%), Cefuroxime (170, 16.3%), Others (81, 7.8%) | 1045 | 53.9 |
| Antibiotics for GPC | Amoxicillin Clavulanate (83, 7.4%), Vancomycin (38, 3.4%), Linezolid (344, 30.0%), Teicoplanin (651, 56.9%), Levonadifloxacin (22, 1.9%)<br>Daptomycin (4, 0.3%) | 1144 | 58.9 |
| Antibiotics for Anaerobes | Clindamycin (138, 45.1%), Metronidazole (168, 54.9%) | 306 | 15.8 |
| Aminoglycosides for GNB | Amikacin (108, 75%), Gentamycin (22, 15.3%), Tobramycin (10, 6.9%), Netilmycin (2, 1.4%), Streptomycin (2, 1.4%) | 144 | 7.4 |
| Quinolones for GNB | Ciprofloxacin (14, 31.1%), Ofloxacin (10, 22.2%), Moxifloxacin (6, 13.3%), Levofloxacin (2, 4.4%), Prulifloxacin (1, 2.2%) | 45 | 2.3 |
| Other Antibiotics for GNB | Trimethoprim-Sulfamethoxazole (50, 55.6%), Nitrofurantoin (38, 42.2%), Chloramphenicol (2, 2.2%) | 90 | 4.6 |
| Other Anti microbials for presumed activity against COVID-19 (many on multiple drugs) | Azithromycin (569, 29.3%), Doxycycline (973, 50.2%), Ivermectin (843, 43.5%) | 1940 | 100.0 |
| Antifungals | Azoles, Echinocandins, Ampho-B | 1105 | 56.9 |
|  | Azoles: Fluconazole (410, 61.4%), Itraconazole (4, 0.6%), Voriconazole (228, 34.1%), Posaconazole (25, 3.7%), Isavuconazole (1, 0.1%) | 668 | 34.4 |
|  | Echinocandins: Caspofungin (58, 29.1%), Anadulafungin (46, 23.1%), Micafungin (95, 47.7%) | 199 | 10.3 |
|  | Amphotericin-B (238, 100%) | 238 | 12.3 |
| Antivirals | Remdesivir (1273, 74.8%), Favipiravir (360, 21.2%), Oseltamivir (40, 2.3%), Lopinavir-Ritonavir (11, 0.6%), Aciclovir (20, 1.1%) | 1704 | 87.8 |

Note: Most of the patients in SI group were on multiple antibiotics and/or antifungals

### Mortality and its Correlates

As shown in Table 5, mortality (40.3%) in the group with SIs was more than 8 times the mortality (4.6%) in the group with no SIs. The proportion of patients getting SIs increased as the severity of COVID-19 increased and so did mortality. In mild COVID-19 group, only 166 patients (2.6%) had SIs, in moderate cases 628 (9.5%), and in severe cases 1146 (16.7%). Mortality in those with SIs with mild, moderate, and severe COVID-19 showed steep increase at 1.2%, 17.5% and 58.5%, respectively (Table 5).

**Table 5:**
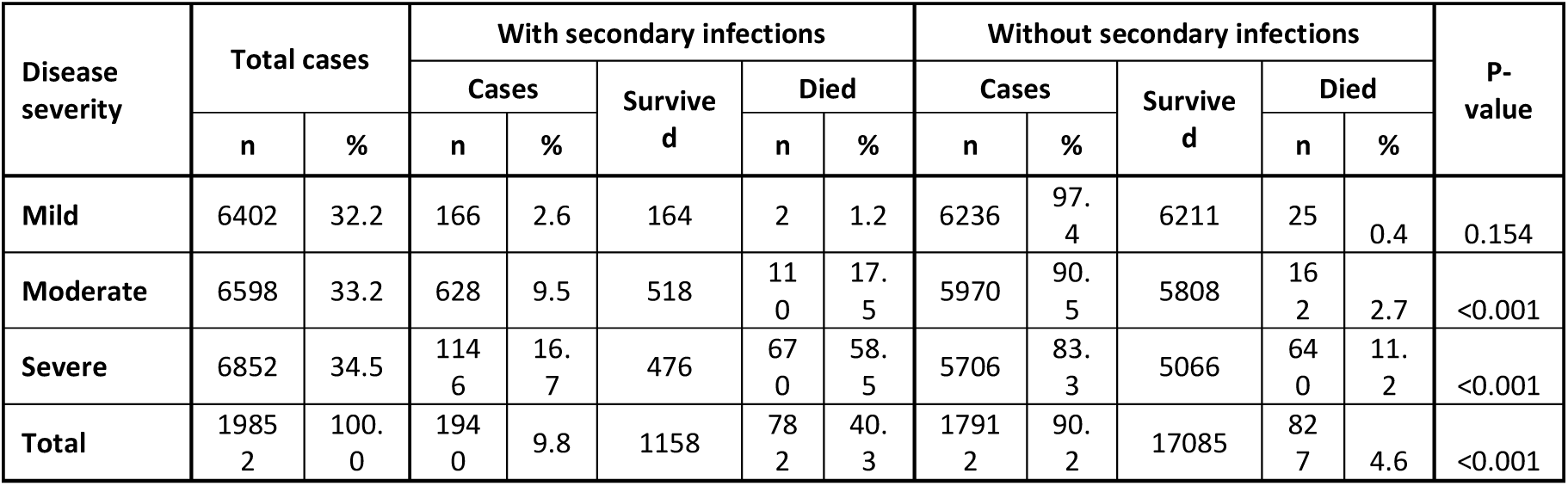
Correlation of mortality with SIs and Severity of COVID-19 disease.

The mortality in relation to the site of infection showed highest mortality (49.8%) in patients with blood stream infection, closely followed by 47.9% with pneumonia, and 29.4% each with urinary and skin / soft tissue infections (Table 6).

**Table 6:**
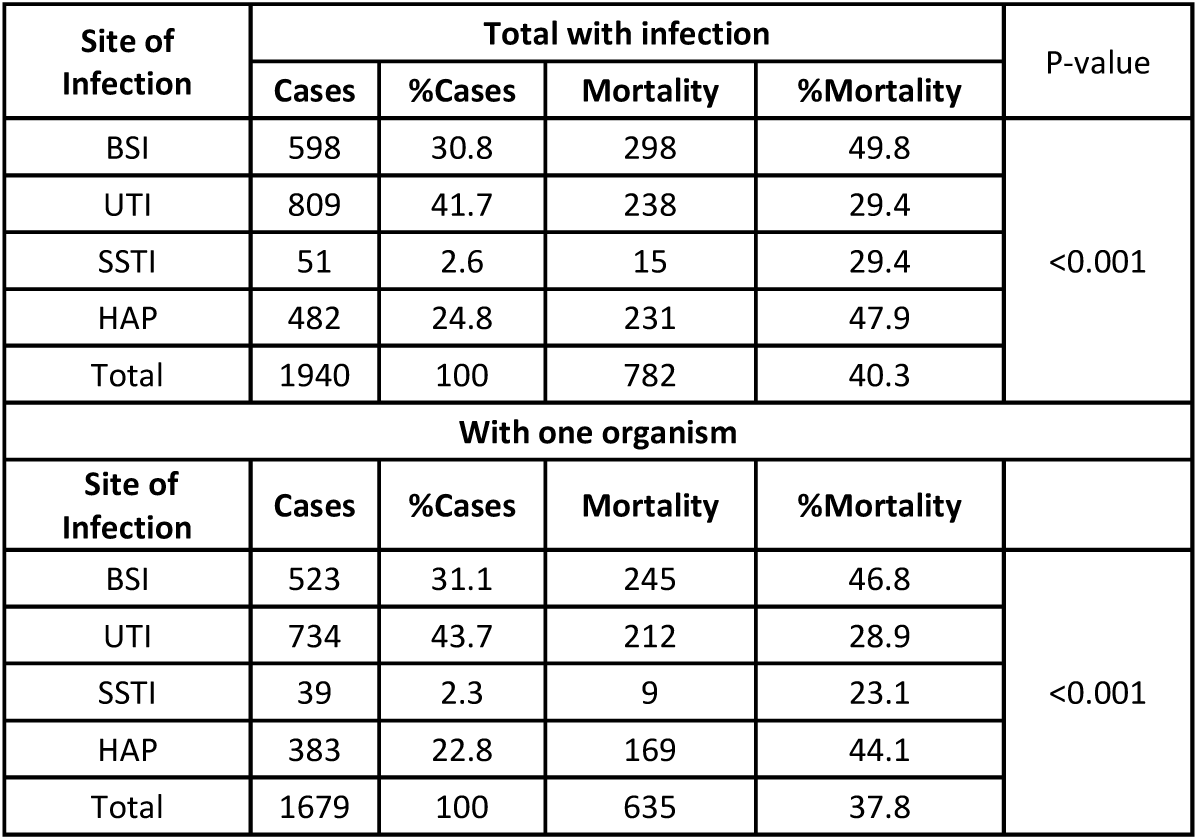
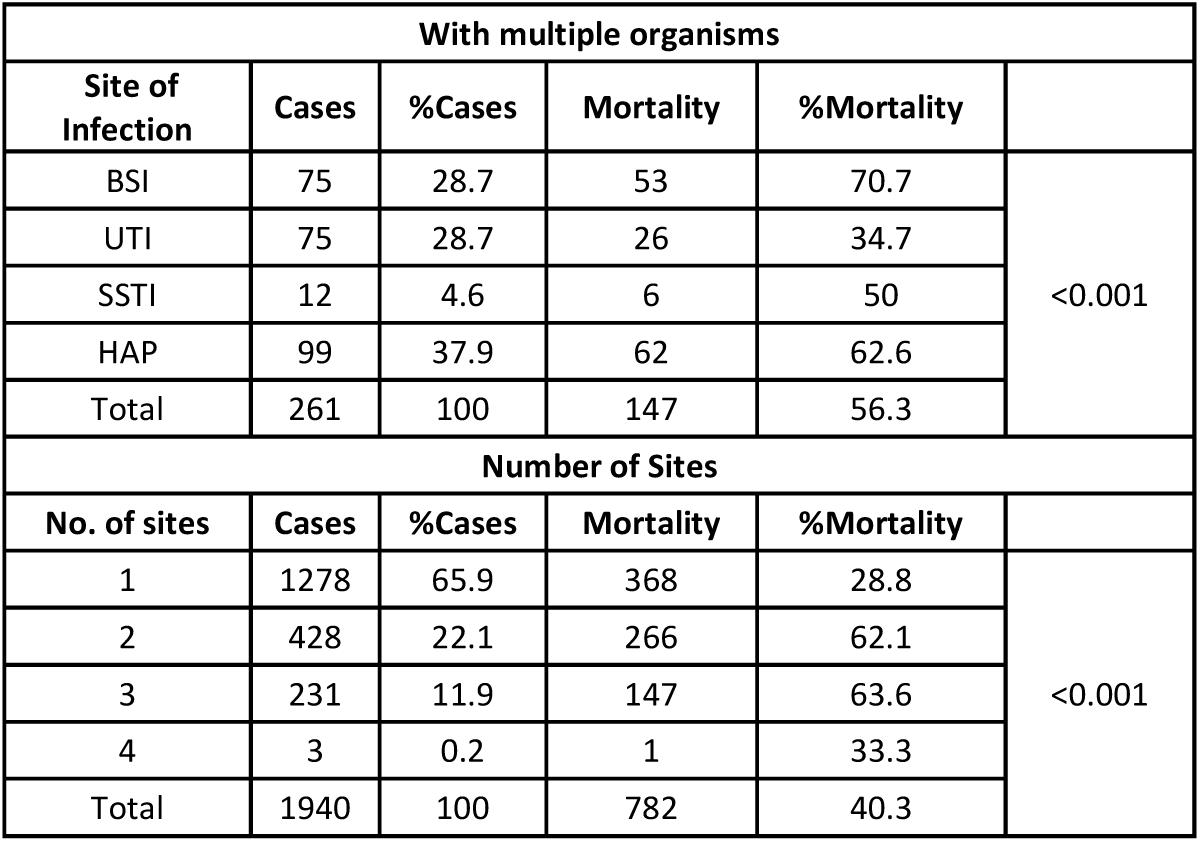
Cases with different sites of infection and their mortality.

The mortality in patients with only one identified organism was 37.8% against 56.3% in patients with more than one organism and was significantly associated with the site of infection in both the cases (p < 0.001). The proportionate pattern of mortality in cases with single and multiple organisms was nearly similar for all sites of infection although the differences were statistically significant (p < 0.001) because of large sample in our study (Table 6). Similarly, mortality in patients with one site of infection was 28.8% against 62.5% in patients with multiple sites of infections (p < 0.001).

More than half (1066, 54.9%) of 1940 who developed SIs were diabetics. The mortality in them was 45.2% against 34.3% in those without diabetes (Table 7) and this difference was statistically significant (p < 0.001).

**Table 7:** Association of mortality with diabetes.

| Diabetes | Cases with SIs | %Cases | Mortality | %Mortality | P-value |
| --- | --- | --- | --- | --- | --- |
| No | 874 | 45.1 | 300 | 34.3 | <0.001 |
| Yes | 1066 | 54.9 | 482 | 45.2 |  |
| Total | 1940 | 100 | 782 | 40.3 |  |

The trend of various inflammatory markers commonly used for monitoring COVID-19 admitted patients showed a higher value in those with SIs versus those without. The median values (Table 8) for CRP, D-dimer, ferritin, IL-6, LDH, and CPK were higher and ALC lower in the group with infection. This difference was even more if we compare the values for those patients who died against those who survived, across both the groups. However, the difference in the median levels of inflammatory markers in those who died in both the groups was not very different and in some markers (such as CRP, IL-6, LDH and CPK) the median values were actually higher in those who had no secondary infection and died. The median CTSS for the overall group with SIs was 15 (IQR: 10-19) and that for the group without SIs was 10 (IQR: 7-14). Again, in those who died in both the groups, the CTSS score was almost similar (median 17) (Table 8).

**Table 8:** Correlation with inflammatory markers.

| LAB Parameters | Total Cases |  |  | Cases with Infections |  |  | Cases without Infections |  |  |
| --- | --- | --- | --- | --- | --- | --- | --- | --- | --- |
|  | All | Survived | Died | All | Survived | Died | All | Survived | Died |
| CRP – n | 13670 | 12633 | 1037 | 1493 | 902 | 591 | 12177 | 11731 | 446 |
| Median | 9.8 | 8.9 | 34.8 | 17.3 | 13.4 | 30.7 | 9.3 | 8.6 | 35.5 |
| (IQR) | (2.3-37.9) | (2.1-33.0) | (10.8-120.5) | (5.5-81.4) | (3.8-52.9) | (8.9-112.3) | (2.1-35.4) | (1.9-32.5) | (12.7-123.8) |
| D-Dimer – n | 13490 | 12434 | 1056 | 1514 | 896 | 618 | 11976 | 11538 | 438 |
| Median | 232.4 | 220.8 | 660.5 | 507.5 | 399.7 | 793.0 | 222.0 | 215.4 | 591.0 |
| (IQR) | (145.7-431.0) | (140.0-381.1) | (318.7-2210.3) | (260.2-1225.8) | (222.0-903.6) | (356.1-2406.8) | (141.0-387.1) | 137.4-363.0) | (306.3-2121.1) |
| Ferritin – n | 11895 | 10940 | 955 | 1350 | 800 | 550 | 10545 | 10140 | 405 |
| Median | 236.6 | 217.8 | 526.8 | 419.6 | 316.6 | 578.6 | 224.0 | 212.4 | 504.0 |
| (IQR) | (102.1- 493.4) | (94.7-455.5) | (270.3-1069.9) | (180.4-841.6) | (140.7-653.8) | (283.3-1068.8) | (97.1-465.0) | (93.1-443.4) | (258.1- |
|  |  |  |  |  |  |  |  |  | 1071.1) |
| IL-6 – n | **11237** | **10218** | **1019** | **1408** | **810** | **598** | **9829** | **9408** | **421** |
| Median | 21.6 | 19.3 |  | 50.1 | 35.4 | 73.2 | 20.0 | 18.7 | 86.5 (33.8- |
| (IQR) | (7.7-56.8) | (7.0-49.0) | 82.6 (31.8-215.4) | (18.8-127.6) | (12.9-86.7) | (31.9-172.5) | (7.2-51.5) | (6.8-46.2) | 232.0) |
| LDH – n | **9773** | **8945** | **828** | **1092** | **629** | **463** | **8681** | **8316** | **365** |
| Median | 301.0 | 291.0 | 517.0 | 410.0 | 345.0 | 517.0 | 295.0 | 288.0 | 519.0 |
| (IQR) | (234.0-402.0) | (230.0-381.0) | (375.8-741.0) | (300.8-573.8) | (261.0-469.0) | (389.0-720.0) | (231.0-390.6) | (229.0-377.0) | (361.5-767.0) |
| CPK – n | **6131** | **5577** | **554** | **648** | **334** | **314** | **5483** | **5243** | **240** |
| Median | 100.0 | 97.0 | 162.0 | 112.5 | 102.5 | 133.0 | 99.0 | 97.0 | 183.0 |
| (IQR) | (59.0-200.0) | (58.0-187.4) | (69.0-370.5) | (55.0-246.0) | (50.0-200.3) | 58.7-316.0) | (59.0-196.0) | (58.0-188.0) | (77.0-444.0) |
| Trop – I – n | **5989** | **5185** | **804** | **998** | **531** | **467** | **4991** | **4654** | **337** |
| Median | 0.0 | 0.0 | 0.0 | 0.0 | 0.0 | 0.1 | 0.0 | 0.0 | 0.0 |
| (IQR) | (0.0-0.0) | (0.0-0.0) | (0.0-0.2) | (0.0-0.1) | (0.0-0.0) | (0.0-0.2) | (0.0-0.0) | (0.0-0.0) | (0.0-0.3) |
| ALC – n | **16620** | **15129** | **1491** | **1883** | **1105** | **778** | **14737** | **14024** | **713** |
| Median | 1.2 | 1.2 | 0.8 | 0.9 | 1.1 | 0.8 | 1.2 | 1.2 | 0.8 |
| (IQR) | (0.8-1.7) | 0.8-1.7) | (0.5-1.3) | (0.6-1.5) | (0.7-1.6) | (0.5-1.3) | (0.8-1.7) | (0.8-1.7) | (0.5-1.2) |
| CTSS – n | **9030** | **8536** | **494** | **847** | **564** | **283** | **8183** | **7972** | **211** |
| Median | 11.0 | 10.0 | 17.0 | 15.0 | 14.0 | 17.0 | 10.0 | 10.0 | 17.0 |
| (IQR) | (7.0-15.0) | (6.0-14.0) | (13.0-21.0) | 10.0-19.0) | (9.0-18.0) | (14.0-21.0) | (7.0-14.0) | (6.0-14.0) | (13.0-21.0) |

## Discussion

COVID-19 patients are at a higher risk of secondary bacterial and fungal infections, and these are associated with increased morbidity and mortality [1,2,3,4]. Several factors are known to contribute to higher risk of secondary infections in these patients. The damage to the respiratory epithelium caused by the virus, as well as their effects on innate and adaptive immunity, antagonising IFN responses that enhance bacterial adherence, colonisation, growth, and invasion into healthy sites in the respiratory tract, are important mechanisms [1,2].Down regulation and differential regulation of immune genes are mechanisms that may create a conducive environment for occurrence of secondary bacterial infections, favouring bacterial attachment to host structural cells and pro-inflammatory environment conducive to suppression of anti-bacterial host defences. In addition, Bogeochea and Bamford [1] suggested SARS-CoV-2-associated perturbation of gut homeostasis as a mechanism that may potentially affect the disease outcomes in patients with severe COVID-19 infection, including predisposing to secondary lung infections.

### Overall Incidence of SIs

Overall, 9.8% of the total hospitalised COVID-19 patients in our network were diagnosed with secondary bacterial or fungal infections. A retrospective study by Vijay et al. [13] on 17,534 COVID-19 patients admitted in 10 hospitals of ICMR-AMR Surveillance network in India, reported SIs in only 3.6% cases. A meta-analysis of 24 studies, including 3338 patients with COVID-19, done by Langford et al. [14] reported overall bacterial infection in 6.9%, and 8.1 % in critically ill patients. Shafran et al. [5] studied 1384 cases (642 COVID-19 cases and 742 influenza cases) for blood and sputum culture results, clinical parameters and outcomes and compared these parameters between the COVID-19 cases and influenza cases. Higher rate of bacterial infection was found in COVID-19 than in those infected with Influenza (12.6% vs. 8.7%). A review of secondary pulmonary infection in patients with COVID-19 pneumonia by Chong et al. [15] from USA reported the incidence of secondary pulmonary infection to be 16% for bacterial infection while 6.3% for fungal infection. Secondary pulmonary infection was predominantly in critically ill hospitalized cases in their study. Thus, the incidence of SIs in COVID-19 patients could be in the range of 5% to 15%, and it increased with the severity of disease.

### Correlates of SIs

We could identify several factors that increased the risk of developing SIs in COVID-19 patients. Average age of patients who had SIs was 8 years more than those without SIs (median 62.6 years versus 54.3 years) (p < 0.001). Also, as age increased, so did the risk of getting SIs. Only 4% of those under 45 years got SIs whereas 18.4% patients above 75 years of age developed SIs. In the earlier Indian study by Vijay et al. the mean age of COVID patients diagnosed with SI was 53.3 ± 9.36 yrs. We observed a higher age of the SI patients and a clear upward gradient of SI incidence with increasing age.

In our present study, 54.9% patients were diabetic in the SIs group. In our another study [16] on similar population base, the overall prevalence of diabetes in COVID-19 admitted patients was 43.8%. This difference is statistically significant (p < 0.001). A study from USA by Adelman et al. [17] reported for their cohort of 774 COVID-19 cases that hypertension was in 75.5% and diabetes mellitus in 45.7% cases. A case-control study conducted in Pakistan by Nasir et al.[18] also reported diabetes mellitus and hypertension as the most common comorbidities in SI cases. In our study, we found that patients with diabetes had relatively more of BSI and UTI and less of SSTI and HAP.

We found a clear correlation between the severity of COVID-19 at the time of admission and the risk of getting SIs. Only 2.6% of those with mild disease developed SIs, while moderate and severe disease had 9.5% and 16.7% respectively. This may be partly related to the need for hospitalisation and ICU stay in the cases with moderate and severe disease. Nasir et al. reported that the critically ill cases at the time of admission were at 4.42 times higher risk for bacterial infection. Chong et al. also found secondary pulmonary infections predominantly in critically ill hospitalized cases. There is a clear relation of higher incidence of SIs with the severity of COVID-19 at admission.

In our study, 22.6% of patients admitted to ICU developed SIs, while only 3% of those admitted in ward got SIs (p < 0.001). Vijay et al. reported that among the cases with confirmed SIs, 71.7% were in ICU while 28.3% were in ward at the time of diagnosis of SI. ICU admission seems to have a definite association with SIs.

It may be difficult to draw a cause-effect relation between the need for oxygen and the risk of getting SIs, but we did observed that SIs developed in 12.1% of patients on oxygen and only 4.9% of those not on oxygen got SIs (P< 0.001). Similarly, the risk of getting SIs increased with increasing need of oxygen and ventilator support. The risk of getting SIs for patients on oxygen by nasal prongs/face mask was 8.4%, for those on NIV was 21.7%, and for those on MV was 23%. This could also be a reflection of more severe disease in those COVID-19 patients who got serious SIs and hence needed to be on a greater support.

### Features of SIs

The most common site of SI in our study was urine (41.7%), followed by blood (41.7%), and pneumonia (24.8%). Skin infection was the least common (2.6%). This spectrum is different from the one reported by Vijay et al. with blood and respiratory as the most common sites of SIs.

Almost 13.4% of the patients had infection with more than one microorganism and 34.1% had multiple sites of infection. Shafran et.al. reported presence of more than 1 coinfection in 4.5% of SARS-CoV-2 cases.

Adelman et al. reported that 30.7% cases required mechanical ventilation and out of these 27.3% had positive respiratory culture with *Staphylococcus aureus* (34.5%) being the most common bacteria followed by *Pseudomonas aeruginosa* (19.0%) and *Klebsiella spp*. (16.7%). Out of 774 cases, blood sample culture was positive in 76% cases and 4.7% (36) had blood stream infection – majority being in ICU (66.7%; 24/36 cases). Shafran et al. reported 85% of isolates to be positive in blood culture while 14.2 % were in respiratory sample. Vijay et al., from India, found overall Gram negative pathogens (78.03%) as the most predominant isolated pathogen, in which most common isolates were *Klebsiella pneumoniae* (29.3%) followed by *Acinetobacter baumanii* (21.07%), *Pseudomonas aeruginosa* (9.6%) and *E. coli* (8.2%). *Candida spp*. were isolated from 6% of admitted cases, of which 1.2% were of *Candida auris. Klebseilla pneumoniae* was the most common isolate in blood (29.7%) and respiratory specimen (35%), while in urine most common isolate was *E*.*coli* (27.17%) followed by *Klebsiella pneumonia* (18.4%) and *Candida spp*. (18.4%). Such varying findings indicate that the spectrum of SIs may be population specific.

### Treatment

We studied the number of patients developing SIs in relation to various treatment modalities used for treatment of COVID-19 patients. This may not necessarily mean that these drugs increased the risk of SIs but may simply reflect high usage of these medicines, especially in sicker patients. Among those patients who received steroids, 10.3% had SIs, while only 3.1% in the group that did not receive steroids. More than one-fourth (26.9%) of those who received convalescent plasma (CP) got SIs, while only 7.3% had SIs in the group that did not receive CP. In the group that got remdesivir, 10.6% had SIs while 8.5% of those who did not receive had SIs. Nasir et al. noted use of systemic steroids to be in significantly higher proportion of cases with bacterial infection than in those without bacterial infection (92% vs. 62%).

Most of our patients with SIs were on multiple antimicrobials. The most used antibiotics were against GNBs. BL-BLI combination therapy was found to be the commonest used treatment (76.9%), followed by carbapenems (57.7%) and cephalosprins (53.9%). Antibiotics directed against GNB-CRE organisms such as Polymyxin B, Colistin, Fosfomycin, Minocycline and Tigicycline were used in 47.1% patients. This matched with the microbiological flora as we identified GNBs to be the cause of SIs in 63.2% cases The high prevalence of CRE in GNBs (68% in *Acinetobacter baumanii*, 48% in *Klebsiella peumoniae*, 39% in *E. coli* and 43% in *Pseudomonas aeruginosa*) justified the empirical usage of antibiotics against CREs. The usage of antibiotics against GPCs (Staphylococcus, CONS, Enterococcus) in our study was found high at 58.9%, whereas the actual microbiological culture data revealed these organisms we identified in only 19.6% samples. Similarly, antifungals were used in 56.9% cases in the present study, while the fungus (Candida sp) was isolated in only 17.3% cases. High degree of azole resistance in various Candida species (54% fluconazole resistance in Candia auris,19% resistance in non-albicans Candida and 10% in *Candida albicans*), and high level of isolation of *Candida auris* (25.7% of Candida isolates in BSI and 13.4% in UTI), *Candida tropicalis* (30.1% in BSI and 45.1% in UTI), justifies the empirical use of Echinocandins (10.3%) and Amphotericin-B (12.3%). However, there is a scope of significant improvement in terms of rationalizing the usage of antimicrobials, especially empiric coverage against GPC and fungus. Each country needs to develop their empiric antibiotic guidelines for hospitalised COVID-19 patients for optimizing the therapy and reduce the potential harm caused by future development of antimicrobial resistance.

Drug resistance profile of the isolated pathogen was studied by Vijay et al. and found 47.1% were infected with multiple drug resistant organisms. – 74.2% of GNB isolates were resistant to carbapenems alone. They reported the use of third generation cephalosporins (16.6%), β-lactam-β-lactamase inhibitors combination (57.3%), and carbapenems (43.7%) in the management of COVID-19 with SI. Vancomycin or teicoplanin was prescribed to 24.9% patients. They also reported that the empirical cover for Gram-positive pathogens may not be warranted as the SIs were predominantly caused by Gram-negative pathogens (78.3%) in their cohort. They also found that 10% patients received antifungals without any evidence of fungal infection. Shafran et al. found that culture reports in cases with either influenza or COVID-19 showed *Pseudomonas aeruginosa* and *Staphyloccocus aureus*as the most common secondary bacterial infections. Gram negative represented 75% of in both groups. Interestingly, enteroccocus infection was found to be more prevalent in cases of COVID-19 than in influenza cases (8.6% vs. 0%), and also late infection with gram positive bacteria was more common in cases with COVID-19 infection. Langford et al. analysed the use of antibiotics in COVID-19 patients and found that over 70% cases received antibiotics, with majority constituted by broad spectrum antibiotics like third generation cephalosporins and fluoroquinoles. Adelman et al. reported that the most common organism isolated were gram negative (28.6%), *Staphylococcus aureus* (16.7%), *Candida species* (16.7%) and *CONS* (11.9%). Nearly 50% were central line associated BSI (CLBSI). Nasir et al. found that among the bacterial infection, gram negative (85%) were more common than gram positive organism. Most frequent organism isolated from blood was MDR *Acinetobacter* followed by *E. coli, Enterococcus*, and *Klebseilla pneumonia*. Among cases with secondary bacterial HAP, the most common isolate was MDR *Acinetobacter* followed by MDR *Pseudomonas aeruginosa*. Chong et al. reported that use of antibiotics was in 60-100% cases in the studies they reviewed, and the most common bacterial microorganism was *Pseudomonas aeruginosa* (21%), followed by *Klebsiella* species (17.2%), *Staphylococcus aureus* (13.5%), *E. coli* (10%), and *Stenotrophomonas maltophilia* (3%). *Aspergillus fumigates* was the most frequently isolated species among fungal infection in COVID-19 cases. Most of the studies showed MDR-GNs to be the commonest organism causing SIs in COVID-19 patients.

### Outcome

The average length of hospitalisation in our SI cases was twice as high as in those without SIs (13 days versus 7 days; p < 0.001). We could identify several risk factors which increased mortality by more than 8 times in SI cases (40.3% vs. 4.6% in the group without SIs). Vijay et al. noted that among cases of COVID-19 with SI, mortality was higher in critically ill patients (68%) compared to the patients in wards (27.6%). Nasir et al. found that cases with COVID-19 having bacterial infection had comparatively greater proportion of deaths compared to controls (42% vs. 18%). The SIs are significant factor for mortality, and they are mostly treatable. Thus, at least some of these deaths can be avoided.

Among the patients who had SIs, severity of COVID-19 disease at the time of admission was correlated with mortality. The mortality in mild, moderate, and severe disease was 1.2%, 17.5% and 58.5%, respectively. The mortality anyways is expected to rise with severity of COVID-19, but SIs may have contributed to the steep rise in gradient. The mortality in the patients with SIs who had diabetes was 45.2% while in the group without diabetes was 34.3% (p < 0.001). The mortality in the group with BSI was highest (49.8%), followed closely in HAP (47.9%) and was 29.4% each in SSTI and UTI group. Adelman et al. found significantly higher overall mortality in COVID-19 cases with BSI compared to those without any BSI (50% vs. 13.8%). However, they did not find any significant difference in mortality rates among the intubated cases with or without identified bacterial respiratory pathogen.

Mortality in our group of patients with SIs, who had only one identifiable microorganism, was 37.8%, which climbed to 56.3% in patients with more than one microorganism (p < 0.001). Shafran et al. reported an overall mortality (in both COVID-19 and influenza cases) of 13.2 % in cases without infection, while mortality was 33% and 61% in cases with one infection and in cases with two infections, respectively. They however, found that in COVID-19 group, mortality was 48.1% in cases with one infection and 75.9% in cases with more than one infection. Patients with SIs at only one site in our series had a mortality of 28.8%, which rose to 62.5% in those with multiple sites of infection (p < 0.001). These finding suggest that secondary infection are significant contributing factor for disease severity among COVID-19 patients leading to higher mortality.

### Laboratory Parameters

The median values for CRP, D-dimer, ferritin, IL-6, LDH and CPK were higher and ALC lower in the group with infection in our study. This difference was even more if we compare the values for those patients who died against those who survived, across both groups. However, the difference in the median levels of inflammatory markers in those who died in both the groups was not very different and for some markers (such as CRP, IL-6, LDH and CPK) the median values were actually higher in those who had no secondary infection and died. Cytokine storm causing significant elevation of these inflammatory markers, independent of secondary infections, would be the most likely reason for this. Nasir et al. found median C-reactive protein (169 vs. 81) and median NLR (8 vs. 4) to be significantly higher in cases with SIs while no significant difference was noted in procalcitonin level (0.36 vs. 0.14) in COVID-19 cases with bacterial infection compared to those without bacterial infection. The median CTSS for the overall group with SIs was 15 (IQR: 10-19) and that for the group without SIs was 10 (IQR: 7-14). Again, in those who died in both the groups, the CTSS score was almost similar (median 17).

## Conclusions

Secondary bacterial and fungal infections can complicate the course of almost 10% of COVID-19 hospitalised patients. These patients tend to not only have a much longer stay in hospital but were also associated with higher requirement for oxygen and ICU care. The mortality in this group rises steeply by as much as 8 times. The group most vulnerable to this complication are those with more severe COVID-19 illness, elderly, and diabetic patients. Judicious empiric use of combination antimicrobials in this set of vulnerable COVID-19 patients can save lives. It seems essential to have a region or country specific guidelines for appropriate use of antibiotics and antifungals to prevent their overuse.

## Data Availability

data of all RTPCR positive COVID19 patients was accessed from Electronic Health Records (EHR) of a network of 10 hospitals across 5 North Indian states, admitted during the period from March 2020 to July 2021.

## Contributions of authors

SB designed the study concept, finalized the draft, and contributed patients for the study, DJ did collation of clinical & lab data, AI did the statistical analysis and contributed to the draft, MA wrote the initial manuscript, BT provided inputs on lab data & interpretation, rest all are clinicians (Internal Medicine, Pulmonology, Critical Care, Lab Medicine and Microbiology) contributed patients/laboratory data and / or treated patients in the present study.

## Acknowledgements

We wish to acknowledge and thank Taruna Sharma for her help at various stages of the study.

